# Pro108Ser mutant of SARS-CoV-2 3CL^pro^ reduces the enzymatic activity and ameliorates COVID-19 severity in Japan

**DOI:** 10.1101/2020.11.24.20235952

**Authors:** Kodai Abe, Yasuaki Kabe, Susumu Uchiyama, Yuka W. Iwasaki, Hirotsugu Ishizu, Yoshifumi Uwamino, Toshiki Takenouchi, Shunsuke Uno, Makoto Ishii, Takahiro Maruno, Masanori Noda, Mitsuru Murata, Naoki Hasegawa, Hideyuki Saya, Yuko Kitagawa, Koichi Fukunaga, Masayuki Amagai, Haruhiko Siomi, Makoto Suematsu, Kenjiro Kosaki, Keio Donner Project

## Abstract

SARS-CoV-2 genome accumulates point mutations constantly. However, whether non-synonymous mutations affect COVID-19 severity through altering viral protein function remains unknown. SARS-CoV-2 genome sequencing revealed that the number of non-synonymous mutations correlated inversely with COVID-19 severity in Tokyo Metropolitan area. Phylogenic tree analyses identified two predominant groups which were differentiated by a set of six-point mutations (four non-synonymous amino acid mutations). Among them, Pro108Ser in 3 chymotrypsin-like protease (3CL^pro^) and Pro151Leu in nucleocapsid protein occurred at conserved locations among β-coronaviruses. Patients with these mutations (N = 48) indicated significantly lower odds ratio for developing hypoxia which required supplemental oxygen (odds ratio 0.24 [95% CI 0.07-0.88, *p*-value = 0.032]) after adjustments for age and sex, versus those lacking this haplotype in the canonical Clade 20B (N = 37). The Pro108Ser 3CL^pro^ enzyme *in vitro* decreases in the activity by 58%, and the hydrogen/deuterium exchange mass spectrometry reveals that mechanisms for reduced activities involve structural perturbation at the substrate-binding region which is positioned behind and distant from the 108th amino acid residue of the enzyme. This mutant strain rapidly outcompeted pre-existing variants to become predominant in Japan. Our results may benefit the efforts underway to design small molecular compounds or antibodies targeting 3CL^pro^.

## Introduction

Severe acute respiratory syndrome coronavirus 2 (SARS-CoV-2) is the cause of the coronavirus disease 2019 (COVID-19) pandemic. As an RNA virus with limited fidelity for genome replication, the SARS-CoV-2 viral genome constantly accumulates point mutations at an average of two nucleotides per months (GISAIDs: http://www.gisaid.org/), while large rearrangements such as a 382-base deletion in the region encoding ORF8 can occur (1,2). Although the 382-base deletion was associated with a milder clinical course, whether the accumulation of point mutations is associated with a better prognosis is unclear.

Our institute, Keio University Hospital, has a catchment area of the Tokyo Metropolitan area and the surrounding prefectures, and we have been performing whole viral genome sequencing of SARS-CoV-2 in COVID-19 patients since March 2020, aiming to characterize healthcare-associated infections rapidly and effectively and to prevent the spread of infection (3). Molecular viral genome sequencing studies have indicated that the number of mutations is indeed increasing (4,5). According to a Japanese governmental report, the daily number of newly identified COVID-19 patients in Tokyo plateaued during the time period when restrictions were imposed on the entry of foreigners to Japan, while the number of seriously ill patients has been decreasing since May 2020 (https://www.mhlw.go.jp/stf/covid-19/kokunainohasseijoukyou.html). Hence, the relative proportion of seriously ill patients has been decreasing, and the mortality rates in Japan appear to be lower than the rates reported in Western countries (https://covid19.who.int/) (6).

We hypothesized that the accumulation of mutations may have contributed to the decrease in clinical virulence. Molecular surveillance of patients infected in the Tokyo Metropolitan area led us to characterize the non-synonymous Pro108Ser mutation in the 3 chymotrypsin-like protease (3CL^pro^) which has been focused on therapeutic targets for feline coronavirus (7).

## Results

### Viral genome sequence analysis

A total of 70 viral haplotypes were observed in the 90 individuals. A mean of 11.8 mutations (SD = 3.1]) separated the lineage from the founding Wuhan haplotype (the central haplotype of clade A). None of the strains had truncating mutations such as frameshift or non-sense mutations. The number counts of non-synonymous mutations among the strains varied from 2 to 12 (mean, 7.5 [SD], 2.4]), as compared with the Wuhan reference strain.

### Clinical background of COVID-19 patients

The clinical characteristics of the 90 patients are shown in Supplementary Table 1. Nineteen patients (23.3%) required supplemental oxygen, and eight (8.9%) developed acute respiratory distress syndrome; five of these eight patients died.

**Table 1:**
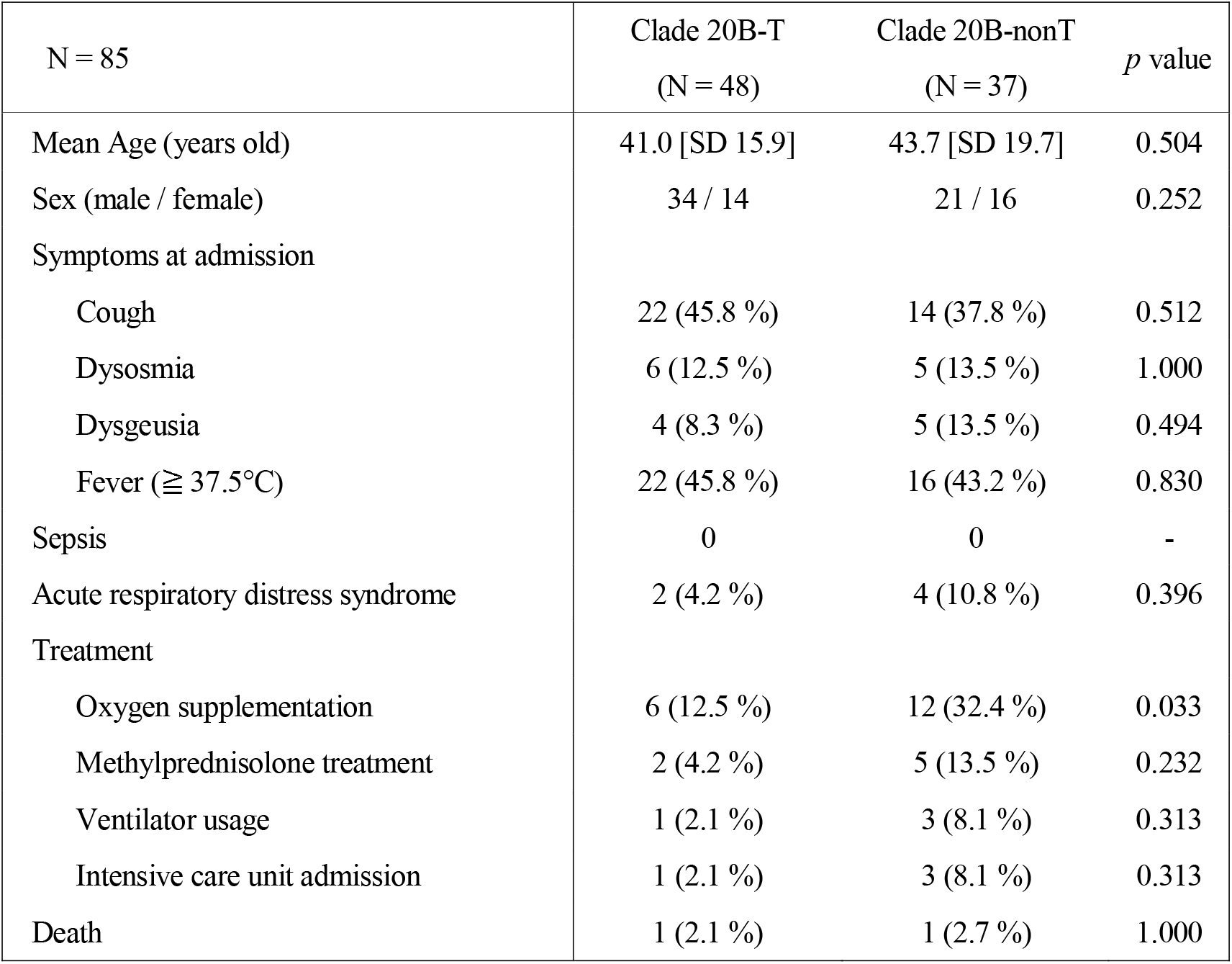
Comparison of clinical features between Clade 20B-T and Clade 20B-nonT.

| N = 85 | Clade 20B-T<br>(N = 48) | Clade 20B-nonT<br>(N = 37) | *p* value |
| --- | --- | --- | --- |
| Mean Age (years old) | 41.0 [SD 15.9] | 43.7 [SD 19.7] | 0.504 |
| Sex (male / female) | 34 / 14 | 21 / 16 | 0.252 |
| Symptoms at admission |  |  |  |
| Cough | 22 (45.8 %) | 14 (37.8 %) | 0.512 |
| Dysosmia | 6 (12.5 %) | 5 (13.5 %) | 1.000 |
| Dysgeusia | 4 (8.3 %) | 5 (13.5 %) | 0.494 |
| Fever ($\geq 37.5^{\circ}\text{C}$) | 22 (45.8 %) | 16 (43.2 %) | 0.830 |
| Sepsis | 0 | 0 | - |
| Acute respiratory distress syndrome | 2 (4.2 %) | 4 (10.8 %) | 0.396 |
| Treatment |  |  |  |
| Oxygen supplementation | 6 (12.5 %) | 12 (32.4 %) | 0.033 |
| Methylprednisolone treatment | 2 (4.2 %) | 5 (13.5 %) | 0.232 |
| Ventilator usage | 1 (2.1 %) | 3 (8.1 %) | 0.313 |
| Intensive care unit admission | 1 (2.1 %) | 3 (8.1 %) | 0.313 |
| Death | 1 (2.1 %) | 1 (2.7 %) | 1.000 |

### Number counts of non-synonymous mutations of SARS-CoV-2 and severity of COVID-19

The number counts of non-synonymous mutations were significantly higher among the COVID-19 patients who did not require supplemental oxygen (Figure 1A: mean, 7.9 [SD 2.2] vs. 5.9 [SD 2.2], *p-*value < 0.001) and increased as the severity decreased (Figure 1B: JT = 404, *p-*value < 0.001). The number counts of non-synonymous mutations with deleterious PROVEAN scores were also higher among the patients who did not require supplemental oxygen (mean, 1.5 [SD, 1.1] vs. 0.9 [SD 0.9], *p*-value = 0.016) and increased as the severity decreased (JT = 556, *p*-value = 0.035).

**Figure 1:**
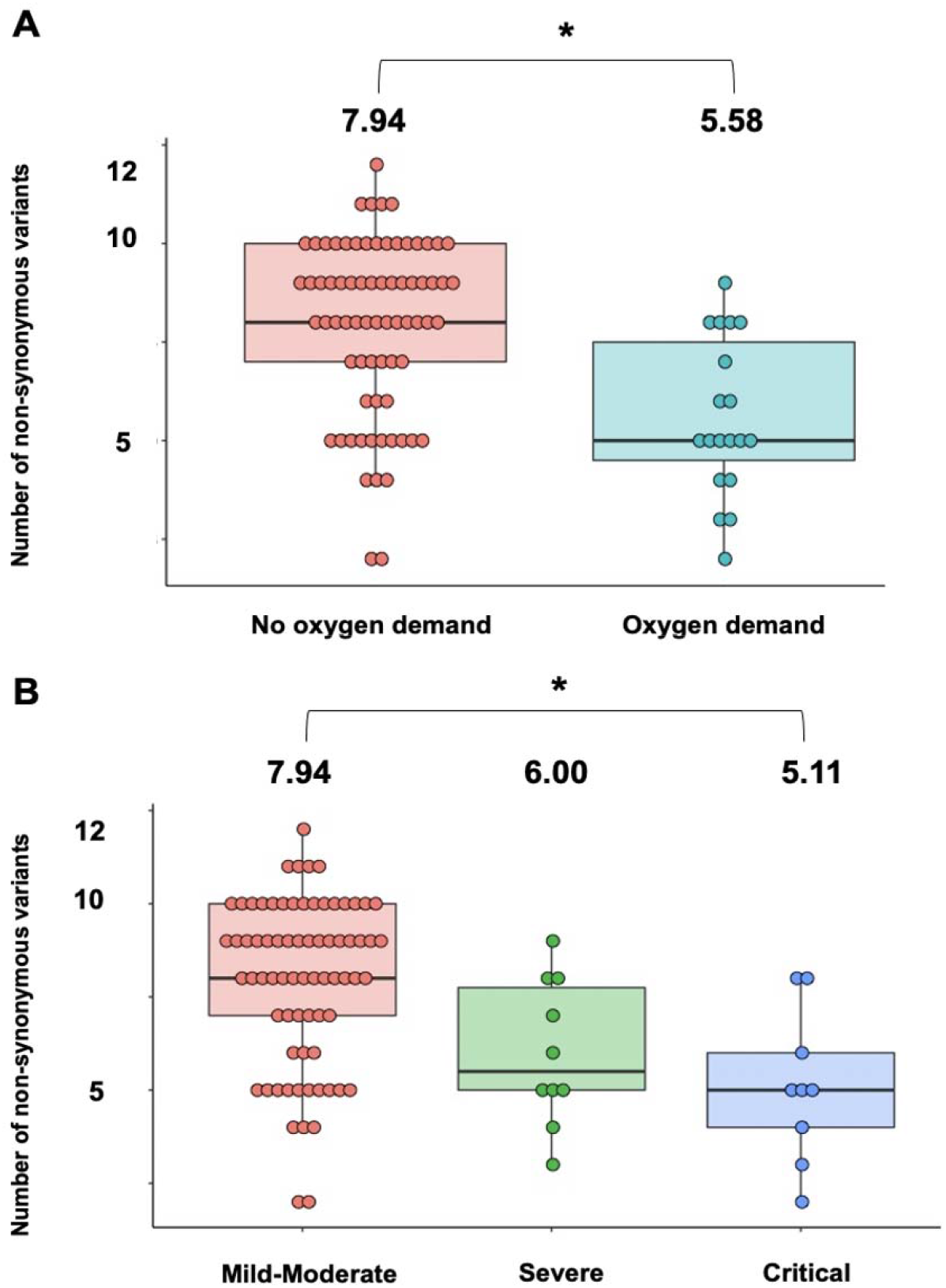
Number of non-synonymous mutations of SARS-CoV-2 is inversely correlated with COVID-19 disease severity. (A) The number of non-synonymous mutations (vertical axis) was significantly lower in COVID-19 patients who required supplemental oxygen than in those who did not (Student’s *t*-test: mean, 7.9 [SD, 2.2] vs. 5.9 [SD, 2.2], \**p*-value < 0.001). (B) The number of non-synonymous mutations (vertical axis) tended to decrease as the COVID-19 severity increased (Jonckheere-Terpstra trend analysis: JT = 404, * *p*-value < 0.001). SARS-CoV-2, severe acute respiratory syndrome coronavirus 2; COVID-19, coronavirus disease 2019; SD, standard deviation; JT, Jonckheere-Terpstra.

### Phylogenic tree analysis

We investigated whether any of the phylogenic clade containing non-synonymous mutations contributed to a milder clinical course. The overall genetic diversity was relatively low, presumably because effective international border restrictions and successful quarantine efforts were in place. A divergent tree analysis of the whole viral genome sequences and classification at Keio University Hospital (N = 90) and in Japan (N = 9106) according to the internationally recommended nomenclature showed that most (i.e., 85 [94.4%] in our study and 8426 [92.5%] in Japan) patients had strains derived from Clade 20B, respectively (Figure 2A) (8). The remaining 5 patients in our cohort study belonged to Clade 19A with the functionally relevant Asp614Gly mutant in the spike protein (9), and were therefore excluded from further study. Patients from Clade 20B (Keio, N = 85; Japan, N = 8426) were additionally divided into two subgroups by defining a subgroup as patients who had strains with no more than 5 nucleotide differences. The first subgroup was designated as the subclade 20B-T (Our study, N = 48 [56.5%]; Japan, N = 4172 [49.5%]), which had the basic haplotype of Clade 20B but had additional 6 single nucleotide mutations: c.4346 U>C, c.9286 C>U, c.10376 C>U, c.14708 C>U, c.28725 C>U and c.29692 C>U (Figure 2A, yellow). Of the six mutations, four were non-synonymous: c.4346 U>C (Ser543Pro in papain-like protease [PL^pro^]), c.10376 C>U (Pro108Ser in 3CL^pro^), c.14708 C>U (Ala423Val in RNA-dependent RNA polymerase [RdRp]), and c.28725 C>U (Pro151Leu in nucleocapsid protein); the remaining two other mutations did not affect the amino acid translation of the viral proteins. The second subgroup was designated as Clade 20B-nonT (Our study, N = 37 [43.5%]; Japan, N = 4254 [50.5%]), which showed the haplotype of Clade 20B, and was defined by seven mutations separating the lineage from the founding Wuhan haplotype, but had fewer than five single nucleotide mutations. Analyses of the cumulative total number and frequency curve showed that the relative fraction of Clade 20B-T increased during the time frame of this study (Figures 2B-C). Mapping of the suspected geographic locations where infection in individual patients took place indicated that the patients with Clade 20B-T or Clade 20B-nonT were infected in the Tokyo Metropolitan area and its neighboring prefectures (Figure 2D). This observation, together with a lack of patients with strains belonging to other clades (except for the 5 patients with Clade 19A who belonged to the same cluster) suggested that Clade 20B and its variation Clade 20B-T were the predominant strains in Japan during the observation period.

**Figure 2:**
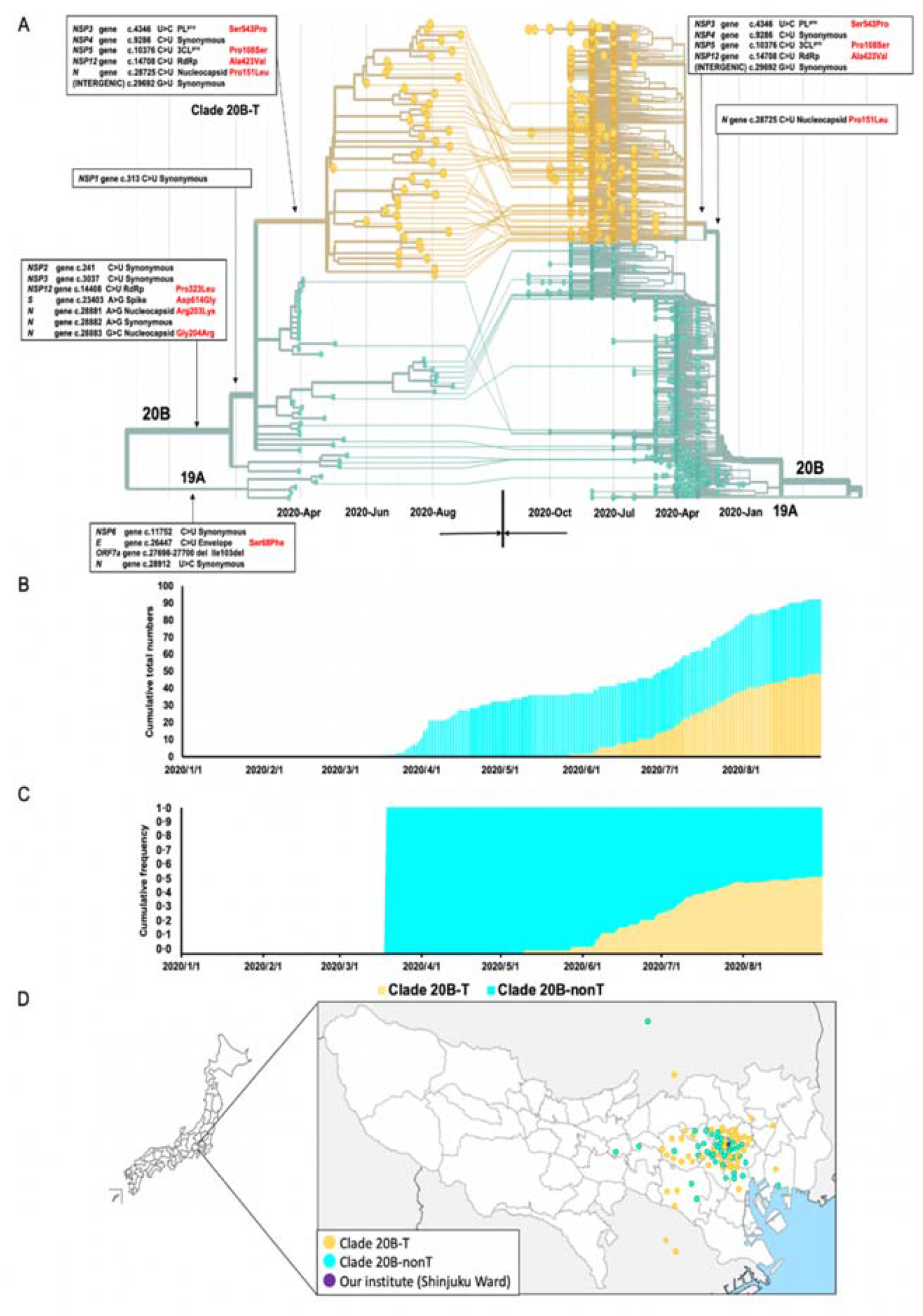
Phylogenic tree analysis, temporal trends, and spatial distribution around Keio University Hospital (purple dot in *Figure* 2D) showed consistent increase of a strain with a unique haplotype. (A) Connection of Keio Strains (right, N = 90) and Japanese strains (left, N = 9106) to the clades defined by GISAID was described in the time-resolved phylogenic tree. Most of the Japanese strains were derived from National Institute of Infectious Diseases (NIID) submitted on December 10 in GISAID, but were not specified by towns/cities or precise obtaining dates (obtaining month only). Therefore, we designated all the NIID data for the first day of the month (i.e., 2020/4 → 2020/4/1). Lines from phylogenic tree of Keio University Hospital (left) to that of all Japanese strains (right) indicate the phylogenetic relation and Nextstrain assignment of Japanese strains. Right- and left-directed arrows in the horizontal axis indicate observation dates in Keio University Hospital and Japan, respectively. Yellow branches represent the predominant Clade “20B-T” both in Keio University Hospital and in Japan which had the six additional mutations compared with the remaining strains Clade “20B-nonT” of Clade 20B. (B) Temporal trends of the number of patients of Clade 20B-T and Clade 20B-nonT at our institute. Clade 20B-T became predominant over Clade 20B-nonT. (C) The cumulative frequency of Clade 20B-T increased steadily to exceed 50% at our institute. (D) The suspected location of infection of individuals from Clade 20B-T and those from Clade 20B-nonT scattered over the Tokyo Metropolitan area and its neighboring prefectures. COVID-19, coronavirus disease 2019; NSP, Non-structural polyprotein; PL^pro^, papain-like proteinase; 3CL^pro^, 3 chymotrypsin-like protease; RdRp, RNA dependent RNA polymerase; ORF, open reading frame.

### Milder clinical course in Clade 20B-T patients

A comparison of the clinical characteristics of the patients with Clade 20B-T (N = 48) with those of the patients with Clade 20B-nonT (N = 37) is shown in Table 1. Age, sex, symptoms at admission, and outcome did not differ significantly between the two main groups, but requiring oxygen supplementation was significantly lower among the patients with Clade 20B-T than in those with Clade 20B-nonT (12.5% vs. 32.4%, *p*-value = 0.033). An exact logistic regression analysis showed that patients with Clade 20B-T had a lower odds ratio for developing hypoxia requiring supplemental oxygen, versus those with Clade 20B-nonT (adjusted odds ratio, 0.24 [95% CI, 0.07-0.88], *p*-value = 0.032; Table 2) after adjustments for age group (< 65 years or ≧65 years) and sex (female or male).

**Table 2:** Logistic regression analysis of candidate predictors for requiring supplemental oxygen.

|  | Univariate model |  | Multivariate model |  |
| --- | --- | --- | --- | --- |
|  | OR (95% CI) | *p* value | Adjusted OR (95% CI) | *p* value |
| Age, years |  |  |  |  |
| < 65 | 1 (ref) | - | 1 (ref) | - |
| ≥ 65 | 12.02 (2.66-65.58) | < 0.001 | 14.67 (3.32-64.79) | < 0.001 |
| Sex |  |  |  |  |
| Female | 1 (ref) | - | - | - |
| Male | 2.20 (0.60-10.19) | 0.269 | - | - |
| Infection |  |  |  |  |
| Clade 20B-nonT | 1 (ref) | - | 1 (ref) | - |
| Clade 20B-T | 0.30 (0.08-1.00) | 0.033 | 0.24 (0.07-0.88) | 0.032 |

OR, odds ratio; CI, confidence interval.

### Molecular evolutionary characterization of four non-synonymous mutations unique to Clade 20B-T

We used molecular evolutionary analyses to decipher which of the four non-synonymous mutations that characterize the Clade 20B-T haplotype contributed to a milder clinical course. Studies of the conservation of the amino acid residues around the non-synonymous mutations in Clade 20B-T indicated that residues at and around Pro108Ser in the 3CL^pro^ (NSP5), and those at and around Pro151Leu in the nucleocapsid protein were highly conserved in *β-*coronaviruses (Figure 3A, B). By contrast, amino acid residues at and around Ser543Pro in the PL^pro^ (NSP3) and Ala423Val in RNA-dependent RNA polymerase (RdRp, NSP12) were only weakly conserved (Figure 3A). On the other hand, the serine in the PL^pro^ at residue 543 and the Ala at residue 423 in RdRp, are substituted with proline and valine in some β-coronaviruses, where both mutations were observed in Clade 20B-T, suggesting that Ser543Pro in the PL^pro^ and Ala423Val in RdRp are likely to be functionally neutral. The PROVEAN predicted that these 2 mutations were not deleterious.

**Figure 3:**
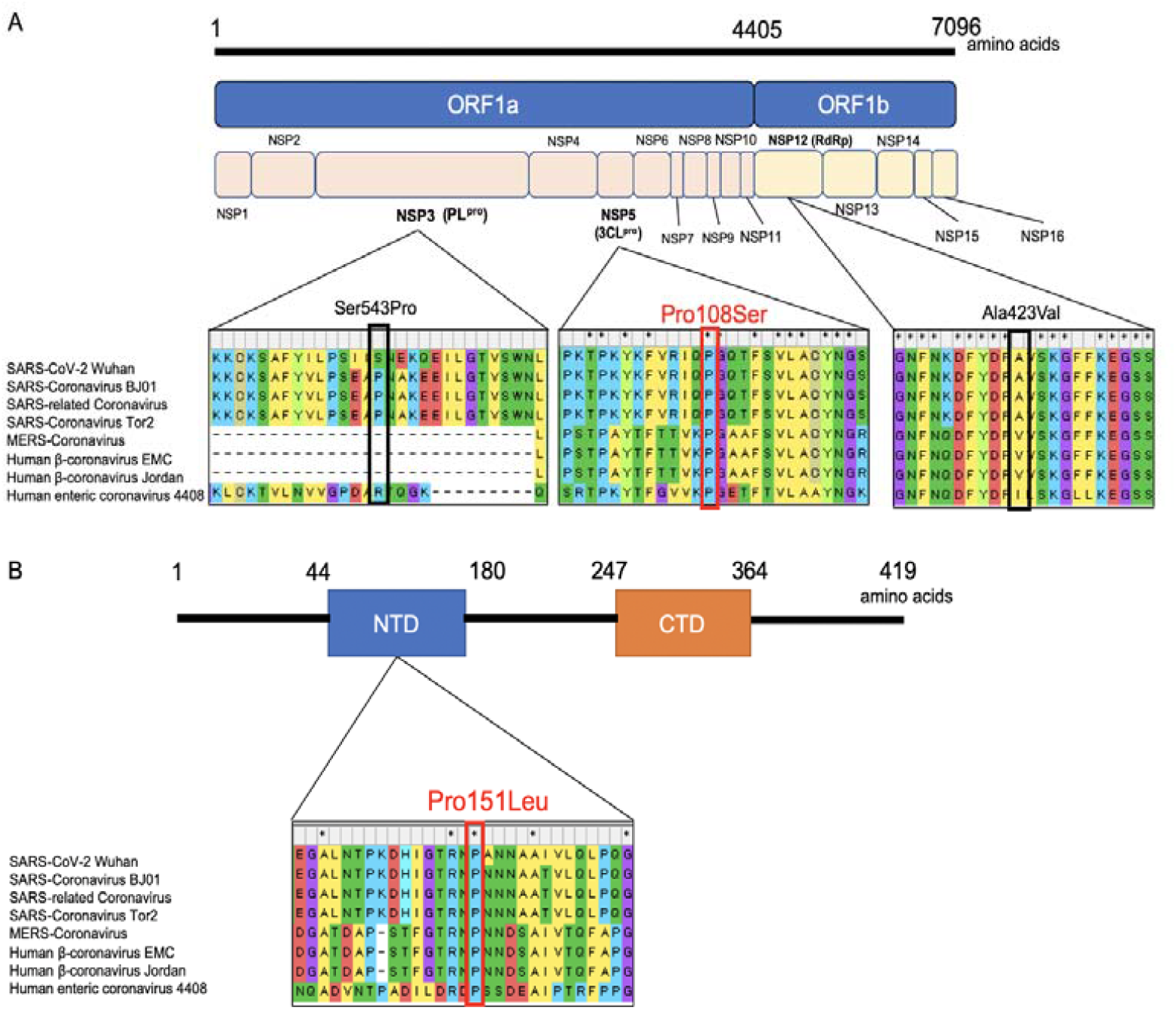
Multiple amino acid sequence alignments of various β-coronaviruses and locations of mutated amino acid residues in Clade 20B-T. (A) The structure of the genomic region that encodes nonstructural polyproteins of SARS-CoV-2. Multiple sequence alignments homologous proteins of 7 β-coronaviruses at and around 3 non-synonymous mutations: Ser543Pro in the PL^pro^, Pro108Ser in the 3CL^pro^, and Ala423Val in the RdRp. (B) The structure of the genomic region that encodes nucleocapsid protein of SARS-CoV-2. Multiple sequence alignments homologous proteins of 7 β-coronaviruses at and around the non-synonymous mutation Pro151Leu in the nucleocapsid protein. SARS-CoV-2, severe acute respiratory syndrome coronavirus 2. ORF, open reading frame; NSP, nonstructural protein; PL^pro^, papain-like protease; 3CL^pro^, 3 chymotrypsin-like protease; RdRp, RNA-dependent RNA polymerase; SARS, severe acute respiratory syndrome; MERS, middle east respiratory syndrome; NTD, N-terminal domain; CTD, C-terminal domain; COVID-19, coronavirus disease 2019; MEGA, Molecular Evolutionary Genetics Analysis.

Thus, Pro108Ser in the 3CL^pro^ and Pro151Leu in the nucleocapsid protein can be considered plausible candidate amino acid mutations that are functionally relevant and may explain a milder clinical course. Because 3CL^pro^ is well characterized by biochemical and pharmacological analysis (10,11) whereas nucleocapsid protein is not, we focused on the function-structure relationship of the P108S mutant of 3CL^pro^ for further investigation.

### P108S 3CL^pro^ reduces the catalytic activity and attenuates the sensitivity to GC376

We prepared recombinant proteins of WT and P108S of SARS-CoV-2 3CL^pro^ to determine the activities by a fluorescence-based cleavage assay (Figure 4A) (11). The enzymatic activity of the P108S was suppressed significantly as compared with that of the WT (Figures 4B). The Km value of P108S (215.7 μM) was lower than that of WT (110.3 μM), and the activity is also decreased by 58%, so far as judged by comparison of the Kcat/Km values of the WT and P108S 3CL^pro^ enzymes (Figure 4C). These results suggest that the P108S mutation interferes with the ability of the enzyme to allow substrate binding.

**Figure 4:**
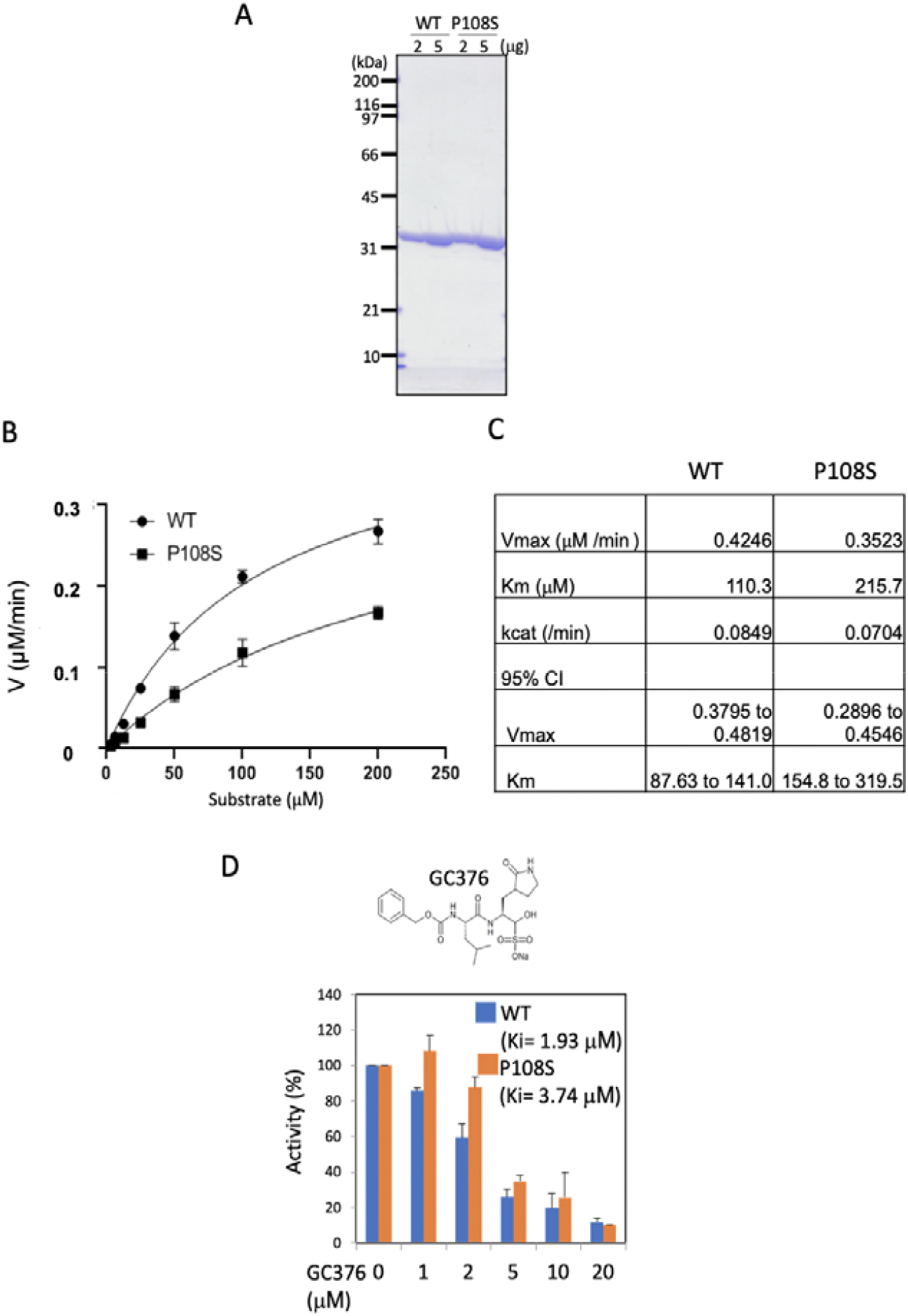
SARS-CoV-2 3CL^pro^ P108S is declined its enzymatic activity by structural alteration. (A) Recombinant WT or P108S of SARS-CoV2 3CL^pro^ were analysed with SDS-PAGE visualizing using CBB staining. (B) The enzymatic activities of SARS-CoV2 3CL^pro^ WT (circle) and P108S (square) were determined using a FRET-based substrate with the cleavage site of SARS CoV-2 3CL^pro^ (Dabcyl-KTSAVLQ↓SGFRKME-Edans). Error bars show mean □ ± □ SD (n=3). (C) The kinetic parameters of enzyme activity of 3CL^pro^ WT and P108S were determined using GraphPad Prism 8 software by initial rate measurement of the substrate cleavage. The Kcat/Km value of the P108S mutant enzyme was 42% of that of the WT enzyme, showing 58% reduction. CI indicates 95% confidence interval. (D) Inhibitory activities of SARS-CoV2 3CL^pro^ WT and P108S by GC376 were analyzed using a FRET-based cleavage assay. The graph shows the relative enzymatic activity. The inhibitory constant (Ki) was calculated using GraphPad Prism 8 software. Error bars show mean□±□SD (n=3). SARS-CoV-2, severe acute respiratory syndrome coronavirus 2; 3CL^pro^, 3 chymotrypsin-like protease; WT, Wuhan-strain type; P108S, Pro108Ser-strain type; SDS-PAGE, Sodium dodecyl sulfate-Polyacrylamide gel electrophoresis; CBB, Coomassie Brilliant Blue; FRET, fluorescence resonance energy transfer.

We further examined the sensitivity of P108S mutant against a competitive 3CL^pro^ inhibitor GC376. Recently, a feline infectious peritonitis virus (FIPV) inhibitor GC376 has been reported to block the SARS-CoV-2 3CL^pro^ activity by binding to the substrate-binding pocket (12,13). The enzymatic activity of WT was potently inhibited (Ki=1.93 μM) by GC376. On the other hand, the inhibitory effect of GC376 on P108S mutant was decreased by Ki= 3.74 μM (Figure 4D).

Since previous studies of SARS-CoV 3CL^pro^ indicates that dimerization of 3CL^pro^ activates its enzymatic activity (14,15), we analyzed the dimeric states of SARS-CoV-2 3CL^pro^ WT and P108S with SV-AUC. Analysis of the WT and P108S similarly showed concentration dependencies of the weight averaged s-value (Supplementary Figure 1), indicating that the values of monomer-dimer dissociation constants are comparable between WT and P108S mutant proteins in the given concentration ranges. The analysis with circular dichroism (CD) spectroscopy showed no discernible differences in the secondary and tertiary structures between the two proteins (Supplementary Figure 2). On the other hand, HDX-MS enabled to detect the structural perturbations around the substrate-binding pocket including C128-L141 close to P108 and Y161-D176 (Figure 5A-D), suggesting that the P108S mutation perturbs the pocket which is behind and distant from the mutation.

**Figure 5:**
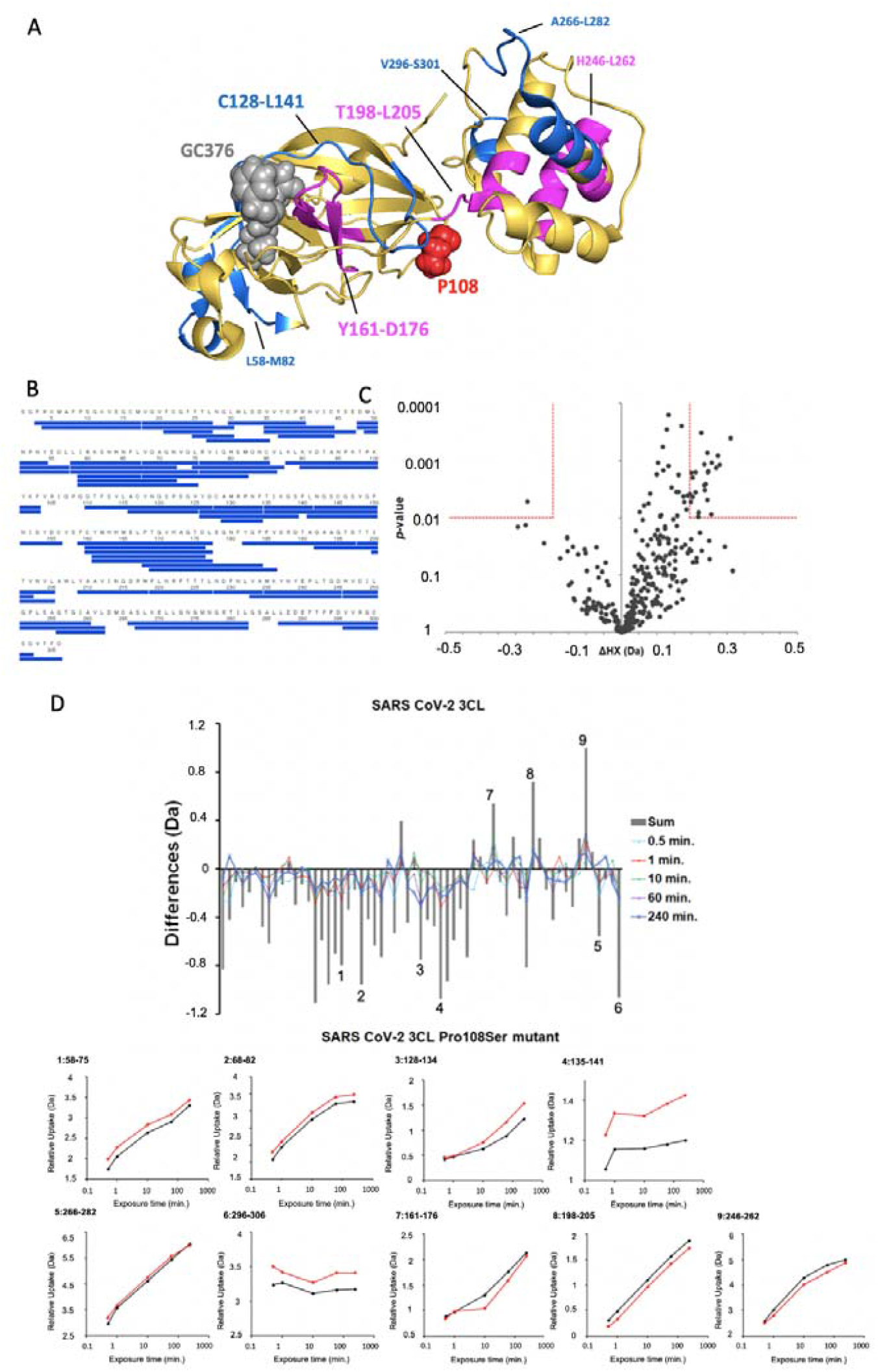
HDX-MS results of SARS CoV-2 3CL^pro^ WT and P108S. (A) Structurally influenced regions accompanied by a single mutation at 108th amino acid from proline to serine. HDX-MS showed more protected regions (magenta) and more exposed regions (cyan) in SARS-CoV-2 3CL^pro^ Pro108Ser mutant compared to SARS-CoV-2 3CL^pro^. Mutation of proline to serine at 108th amino acid induces structural alternation at the regions from C128 to L141 and from Y161 to D176, where C128-L141 is sandwiched between P108 and Y161-D176 which is located at the substrate binding region. (B) The coverage map of identified peptides in SARS-CoV-2 3CL^pro^. (C) Volcano plots of observed delta HDX values and *p*-values calculated from Welch’s t-test for SARS-CoV-2 3CL WT. Red lines showed the horizontal *p*-value and the vertical delta HDX values for the significant criteria. (D) Differential plots of deuterium uptake degrees of peptides, showing time courses, along with their summational results (gray bar). Deuterium uptake curves for the peptides showing significant differences between WT (black) and P108S (red) proteins were presented. SARS-CoV-2, severe acute respiratory syndrome coronavirus 2; 3CL^pro^, 3 chymotrypsin-like protease; WT, Wuhan-strain type; P108S, Pro108Ser mutant. HDX-MS, Hydrogen/deuterium exchange mass spectrometry.

## Discussion

The number count of non-synonymous mutations was inversely correlated with the severity of COVID-19 disease in a cohort of 90 patients whose viral genome sequences had been completely sequenced. Patients with a viral haplotype containing the 3CL^pro^ Pro108Ser tended to have a milder disease course than those with a viral haplotype lacking the mutation. Furthermore, the Kcat/Km value of 3CL^pro^ containing Ser108 was decreased by 58% as compared with that of 3CL^pro^ containing Pro108. Altogether, these clinical and experimental observation suggest that Pro108Ser, which is becoming prevalent in Tokyo, is associated with milder clinical course.

3CL^pro^ is the main protease that cleaves viral polyproteins into functional proteins (16). Based on the SV-AUC, CD spectroscopy, and HDX-MS results, it can be concluded that although the mutation of Pro108 to Ser does not affect the overall structure or association state of 3CL^pro^, the substrate binding site is locally impacted by the mutation, leading to the reduced binding affinity to substrate rather than the monomer-dimer transition of the protease. Thus, Pro108Ser may play an important role in viral replication and pathogenicity.

Since the time of the SARS-CoV epidemic in 2002 and 2003, various therapeutic agents and vaccines have been developed, but not with particularly satisfactory results (17). Apart from human coronavirus infections, GC376 has been shown to be effective against FIPV, which belongs to the α-coronavirus family. GC376 exerts its antiviral action through a reduction in viral replication by creating a covalent bond with the active sites in 3CL^pro^ protein (18,19). The administration of GC376 is associated with a high rate of disease remission and no significant side effects when used against FIPV (20). These findings together with recent studies showing an inhibitory effect of GC376 on the 3CL^pro^ of SARS-CoV-2 through the repression of viral replication *in vitro* suggest that GC376 and its homologs represent promising therapeutic options (7,12,13). Although Pro108Ser decreased the activity of GC376, both Pro108Ser and GC376 had the inhibitory effects on 3CL^pro^, giving an important clue to developing an anti-viral drug targeting 3CL^pro^ (21).

In general, RNA viruses continue to survive and proliferate by constantly changing their forms through mutations and adapting to various environments, but quasispecies with high replicability have difficulty when it comes to long-term survival because of their high sensitivity to environmental influences (i.e., survival of the fittest) (22). The latest observation in the United Kingdom that the strain VOC 202012/01 emerged in southeast England in November 2020 has rapidly spread towards fixation is compatible with this theory (23). On the other hand, quasispecies with low replicability may have reduced infectivity and pathogenicity but actually have long-term survival advantages because they are less likely to be subjected to natural selection (i.e., survival of the flattest) (24). Coronaviruses, including SARS-CoV or SARS-CoV-2, are known to have relatively high but incomplete fidelity in replicating their genome (25). Of the proteins involved in the replication of the viral genome, proteases, including 3CL^pro^, are critical for viral replication. Therefore, Pro108Ser in 3CL^pro^ may lead to a situation in which the virus is less susceptible to natural selection and better suited to long-term survival, reducing viral infectivity and virulence. In fact, Japan whole genome data (N = 9106) recently updated on 10^th^ December 2020 revealed that the Japanese-specific Clade 20B-T including Pro108Ser in 3CL^pro^ progressed to fixation within the Clade 20B during the time period when restrictions were imposed on the entry of foreigners to Japan. We should observe carefully whether Clade 20B-T with these unique mutations would represent a survival of the flattest in Japan.

The present study was limitated in that we could not fully explore the contribution of the other putatively deleterious mutation Pro151Leu in tetramer-forming nucleocapsid protein, which enters the host cells along with the viral RNA and is responsible for processing the assembly and release of viral particles (26). The computer-based protein structure predicted that the Pro151Leu mutation in nucleocapsid proteins is less flexible, but has no significant changes in protein structure or stabilities (Supplementary Figure 3).

In conclusion, viral genome sequencing in Tokyo showed that Pro108Ser in 3CL^pro^, ameliorates the COVID-19 severity. The specific mutant strain containing Pro108Ser in 3CL^pro^ constitutes a major Clade 20B-Tokyo. Our protein analysis demonstrated that the 3CL^pro^ mutant reduces the function of the protein. The mutant strain rapidly outcompeted pre-existing variants to become the dominant one in Japan. Our results may benefit the efforts under way to design small molecular compounds or antibodies targeting 3CL^pro^.

## Methods

### Study design and patients

A total of 187 patients who had been diagnosed with COVID-19 between 17^th^ March and 31^st^ August on the basis of reverse transcription polymerase chain reaction (RT-PCR) results at Keio University Hospital were enrolled. Of these 187 patients, 134 (71.7 %) underwent whole viral genome sequencing. Of these, 44 patients with only partial genome sequences resulting from insufficient PCR amplification were excluded, leaving 90 patients for analysis (Supplementary Figure 4). Thirty-two of these 90 patients had been reported previously (3). The medical records of all 90 patients were reviewed to obtain data on clinical characteristics and treatments received, and PCR data obtained from samples collected from the nasopharynx, sputum or saliva were collected. The study protocol was approved by the Ethics Committee of Keio University School of Medicine (approval number: 20200062) and was conducted according to the principles of the Declaration of Helsinki.

### Definitions and classification of disease severity of COVID-19 infection

Disease severity of patients was classified according to the clinical management guidelines of the World Health Organization (27) and Japan’s Ministry of Health, Labour, and Welfare (https://www.mhlw.go.jp/content/000650160.pdf). In some of patients with mild or moderate symptoms, the presence of pneumonia could not be determined because they did not have chest X-ray or computed tomography examinations. Therefore, we classified disease severity into the following three categories: “mild to moderate,” (patients did not require supplementary oxygen); “severe,” (patients required oxygen supplementation but not a ventilator); and “critical,” (patients who developed sepsis or acute respiratory distress syndrome and required a ventilator [Supplementary Table 2]) (27).

### DNA sequencing method

The whole viral genome sequences, PCR-based amplification and phylogenic tree analysis were determined as previously reported (Takenouchi et al.) (3). All point mutations including non-synonymous and synonymous mutations were annotated with ANNOVAR software and assessed with VarSifter (https://research.nhgri.nih.gov/software/VarSifter/). The multiple amino acid sequence alignments of various β-coronaviruses were compared with Molecular Evolutionary Genetic Analysis software (MEGA, https://www.megasoftware.net/) (Supplementary Table 3). The functional relevance of non-synonymous mutations was predicted with a Protein Variation Effect Analyzer (PROVEAN v1.1.3, http://provean.jcvi.org/seq_submit.php), the calculations of which are not dependent on sequence conservation among animals. Scores under a threshold value of −2.50 were considered deleterious.

### Cloning and protein preparation of SARS-CoV-2 3CL^pro^

The SARS-CoV-2 3CL^pro^ DNA fragments encoding the Wuhan strain or the strain containing Pro108Ser in *non-structural polyprotein 5* (*NSP5)* gene were prepared using a reverse transcription kit (SuperScript III, ThermoFishher) and were amplified by PCR using primers (forward: 5’-TTTGGATCCAGTGGTTTTAGAAAAATGGCA-3’, reverse: 5’-TTTGTCGACTCATTGGAAAGTAACACCTGAGCA-3’). The fragments were digested with Bam HI and Sal I and then ligated into pCold GST containing the cleavage site for PreScision Protease (GE Healthcare) at the N-terminal region. The expression vectors for the 3CL^pro^ Wuhan strain type (WT) or the Pro108Ser mutant (P108S) were transformed into BL21 (DE3), and the bacteria were incubated in LB with ampicillin at 37°C until OD600 was reached at 0.8. Protein expression was induced by 1 mM isopropyl-β-thiogalactopyranoside for 16 h at 4°C. The cell pellets were re-suspended in a buffer containing 20 mM Tris-HCl (pH7.5), 100 mM NaCl, and 0.1% Tween 20, sonicated twice for 5 min at 4°C, and centrifuged at 20,000 × *g* for 30 min. The supernatant was incubated with glutathione Sepharose 4B (GE Healthcare) for 2h at 4°C. The resin was then washed five times with the same buffer, and the GST tag was cleared by the addition of PreScision Protease and further incubation for 16 h at 4°C. Then, the 3CL^pro^ was prepared using size-exclusion chromatography (Superdex 200; GE Healthcare).

### Enzyme kinetics analysis using fluorescence resonance energy transfer-based assay

The enzymatic activities of 3CL^pro^ WT and P108S were determined using a fluorescent substrate with the cleavage site of SARS CoV-2 3CL^pro^ (Dabcyl-KTSAVLQ↓SGFRKME-Edans; GL Biochem). 3CL^pro^ WT or P108S at a final concentration of 5 μM was incubated in a buffer of 20 mM Tris-HCl (pH7.5), 100 mM NaCl, and 5 mM DTT with the addition of the substrate at a final concentration of 3.125, 6.25, 12.5, 25, 50, 100 or 200 μM at room temperature. The change in fluorescence intensity was monitored with a fluorescence spectrophotometer (Cytation 5; BioTek) at an emission wavelength of 460 nm and an excitation wavelength at 340 nm. The kinetic parameters were determined with GraphPad Prism 8 software and the initial rate measurement of the substrate cleavage. For the inhibition assay, the SARS-CoV 3CL^pro^ inhibitor GC376 (Selleck) at a final concentration of 1, 2, 5, 10, or 20 μM was incubated with 5 μM 3CL^pro^ WT or P108S and 12.5, 25 or 50 μM of substrate.

### Circular dichroism (CD) spectroscopy

CD spectra were collected in the far-UV (200-260 nm) and the near-UV (250-340 nm) spectral regions. Spectra were recorded with a CD spectrometer J-1500 (JASCO Corporation) in a quartz cuvette (1 mm cell length for far-UV and 10 mm for near-UV) at 20°C. The protein samples were prepared in 20 mM Tris-HCl buffer solution (pH 7.3) containing 150 mM NaCl with concentration of 5 µM for the far-UV CD measurements and 20 µM for the near-UV CD measurements. The spectrum of the buffer was measured as a blank and was subtracted from the sample data. Four scans were averaged for each spectral region with a scan rate of 50 nm/min. The data pitch and the bandwidth were 0.5 nm and 1 nm, respectively.

### Sedimentation velocity analytical ultracentrifugation (SV-AUC)

SV-AUC experiments were performed using the Optima AUC (Beckman Coulter) at 20°C with 1, 2.5, 5, 10, 20, 40, and 80 µM of 3CL^pro^ WT and P108S dissolved in 20 mM Tris-HCl buffer solution (pH7.3) containing 150 mM NaCl. Next, 390 µL of each sample was loaded into the sample sector of a 12-mm double-sector charcoal-filled Epon centerpiece, and 400 µl of buffer was loaded into the reference sector of each cell. Data collection was performed at 42,000 rpm using a UV detection system. Data were collected every 240 s with a radial increment of 10 µm at 230 nm for 1, 2.5, and 5 µM samples, at 235 nm for 10 µM samples, at 240 nm for 20 µM samples, at 290 nm for 40 µM samples, and at 295 nm for 80 µM samples. The collected data were analyzed using a continuous *c*(*s*) distribution model implemented in program SEDFIT (version 16.2b) (28) with fitting for the frictional ratio, meniscus, time-inmutation noise, and radial-inmutation noise. Both of the partial specific volumes of WT 3CL^pro^ and the P108S were 0.731 cm^3^/g, which was calculated based on the amino acid composition of each sample using the program SEDNTERP 1.09. The buffer density and viscosity were calculated using program SEDNTERP 1.09 as 1.00499 g/cm^3^ and 1.0214 cP, respectively. Figures of the *c* (*s*_20, w_) distribution were generated using the program GUSSI (version 1.3.2) (29). The weight-average sedimentation coefficient of each sample was calculated by integrating the range of sedimentation coefficients where peaks with an obvious concentration dependence were observed. To determine the dissociation constant of the monomer-dimer equilibrium (KD), the concentration dependence of the weight-average sedimentation coefficient was fitted to the monomer-dimer self-association model implemented in the program SEDPHAT (version 15.2b) (30,31).

### Hydrogen Deuterium Exchange Mass Spectrometry (HDX-MS)

HDX-MS experiments were conducted using Waters HDX with LEAP system (Waters). 80 μM protein solutions (SARS CoV-2 3CL^pro^ WT and SARS CoV-2 3CL^pro^ P108S) were diluted 20-fold with 20 mM Tris-HCl buffer solution (pD 7.3) prepared with D_2_O containing 150 mM NaCl, and incubated at 20°C for various hydrogen/deuterium exchange time period (0.5, 1, 10, 60, or 240 min). The concentration of the protein solution during deuterium exchange was 4 μM, and based on the *K*_D_ estimated from SV-AUC, each protein was considered to be present in more than 98% monomer. The exchange reaction was quenched by dropping the pH to 2.4 with mixing equal volume of 4 M Guanidinium chloride, 0.5 M Tris (2-carboxyethyl) phosphine hydrochloride (TCEP), pH 2.2. One hundred pmol of quenched samples were immediately injected, desalted, and separated online using a Waters UPLC system based on a nanoACQUITY platform. The online digestion was performed over 5 min in water containing 0.05% formic acid at 4°C at a flow rate of 100 μl/min. The digested peptides were trapped on an ACQUITY UPLC BEH C18 1.7 μm peptide trap (Waters) maintained at 0°C and desalted with water and 0.1% formic acid. Flow was diverted by a switching valve, and the trapped peptide fragments were eluted at 40 μl/min onto a column of 1 × 100 mm (C18 1.7 μm, ACQUITY UPLC BEH, Waters) held at 0°C, with a 12 min linear acetonitrile gradient (8–40%) containing 0.1% formic acid. The eluate was directed into a mass spectrometer (Synapt HD, Waters) with electrospray ionization and lock mass correction (using Glu-fibrinogen peptide B). Mass spectra were transformed using MassLynx (Waters) and acquired over the *m/z* range of 100–2000. Pepsin fragments were identified using a combination of exact mass and MS/MS, aided by ProteinLynx Global SERVER (PLGS, Waters). Peptide deuterium levels were determined using DynamX 3.0 (Waters). The relative deuterium uptake percentage was calculated for each peptide by dividing the mean of deuterium uptake, 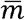, by the number of backbone amide hydrogens. In comparing the HDX results between two samples, the mass difference of hydrogen deuterium exchange for each peptide at each exposure time point (Δ*HX*) was calculated as follow;

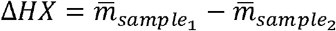

For the statistical analysis of significant difference, the volcano plot, which is a scatter-plot of Δ*HX* versus a probability value (*p*-value) determined from Welch’s t-test, was used (32). Significance limits for vertical Δ*HX* value was calculated as follow. A pooled sample standard deviation (*s*_*p*_) for 610 standard deviations was calculated from Δ*HX*. A propagated standard error of the mean (SEM_Δ*HX*_) was calculated from *s*_*p*_. A significance limit for Δ*HX* values can be calculated by the following equation;

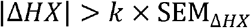

We set *k* = 4.60 by a Student’s t-distribution value for a two-tailed test with four degrees of freedom at a significance level (α) of 0.01 (99% confidence level). For horizontal *p*-value, significance limits were defined at α = 0.01 (99% confidence level).

### Protein structure modeling and stabilization analysis

The three-dimensional (3D) structure was visualized using PyMol v2.4. (https://pymol.org/2/) based on publicly available SARS-CoV-2 protein structure coordinate for the tetramer of nucleocapsid protein (protein data bank ID “6VYO”). We then used the DynaMut server (http://biosig.unimelb.edu.au/dynamut/) to infer the effect of non-synonymous mutations on the nucleocapsid protein 3D structures in terms of molecular stabilities, and flexibilities (33). We also estimated the differences of free energy change (ΔΔ*G*) and vibrational entropy change (ΔΔ*S*Vib) accompanied by the mutation using DynaMut (34), which implements ENCoM reports on the impact on protein stability and flexibility accompanied by mutations in the wild-type structure. Structural changes, such as changes in cavity volume, packing density, and accessible surface area, are correlated with ΔΔ*G*, thereby ΔΔ*G* can be used as an indicator of the impact of a mutation on protein stability (35). A ΔΔ*G* value of less than zero indicates that the mutation causes destabilization, while a ΔΔ*G* above zero indicates protein stabilization.

### Statistical analysis

The main parameter of the clinical study was the grade of disease severity. Comparisons of categorical variables between two groups were assessed using the Fisher exact test. A Student’s *t*-test was used to compare abnormally distributed quantitative variables between two groups, and a Jonckheere-Terpstra test was used to analyze the tendency among three groups. An exact logistic regression analysis was used to examine the odds ratio for requiring supplemental oxygen. The following covariates were considered for inclusion in the multivariate model: age group (< 65 years or ≧ 65 years), sex, and infection group (Clade 20B-T or 20B-nonT). Statistical analyses were performed using R statistical Software (version 3.6.2), and all statistical tests were two-sided. *p-*values < 0.05 were considered significant.

## Supporting information

Acknowledgment table

## Data Availability

We downloaded the full nucleotide sequences of the SARS-CoV-2 genomes from the GISAID database (https://www.gisaid.org/). A table of the contributors is available (acknowledgment table). We have uploaded the full nucleotide sequences of our cohort to the GISAID database.

## Author contributions

KA contributed to writing of the report and data analysis. YK, SUc, TM, MN and MS contributed to review and editing of the report and data analysis for enzyme assay and structural analyses of the 3CL^pro^ proteins. YI, HI, TT and HS contributed to sequencing and analysis. YU, SU and NH contributed to public health intelligence and case identification. MI and KF contributed to clinical data and clinical care. HS, YK and MA contributed to writing and editing of the report. MM contributed to diagnostics and laboratory management. MS and KK had the original idea for the study and contributed to diagnostics, formal analysis, and writing and editing of the report.

## Acknowledgements

The authors thank Professor Timothy Minton for careful review of the manuscript. We thank all the patients and healthcare workers who have fought against COVID-19. This work was supported by the Keio Donner Project and is devoted to the late Professor Shibasaburo Kitasato, the founder of Keio University School of Medicine. We also thank SUNTORY Co., Ltd.

## Data sharing

We downloaded the full nucleotide sequences of the SARS-CoV-2 genomes from the GISAID database (https://www.gisaid.org/). A table of the contributors is available (acknowledgment table). We have uploaded the full nucleotide sequences of our cohort to the GISAID database. This preprint is a new version of our previous preprint which was open in Nov 24, 2020 in medRxiv (doi: https://doi.org/10.1101/2020.11.24.20235952).

