## Supplementary information for "Pro108Ser mutant of SARS-CoV-2 3CL^pro^ reduces the enzymatic activity and ameliorates COVID-19 severity in Japan"

### ***Supplementary Table 1:* Clinical backgrounds of COVID-19 included 90 patients**

|  | N= 90 |
| --- | --- |
| Mean age (years old) | 43.1 [SD 18.3] |
| Sex (male / female) | 58 / 32 |
| Symptoms at admission |  |
| Cough | 38 (42.2 %) |
| Dysosmia | 11 (12.2 %) |
| dysgeusia | 10 (11.1 %) |
| Fever (≧ 37.5°C) | 40 (44.4 %) |
| Sepsis | 1 (1.1 %) |
| Acute respiratory distress syndrome | 8 (8.9 %) |
| Treatment |  |
| Oxygen administration | 21 (23.3 %) |
| Methylprednisolone treatment | 10 (11.1 %) |
| Ventilator | 6 (6.7 %) |
| Intensive care unit admission | 6 (6.7 %) |
| Death | 5 (5.6 %) |
| COVID-19, coronavirus disease 2019. | |

### ***Supplementary Table 2:* Severity of COVID-19 patients**

| Grade | Definition | Total number |
| --- | --- | --- |
| Critical | Patients who developed sepsis or acute respiratory distress syndrome with need for ventilators | 9 |
| Severe | Patients who needed oxygen administration with no need for ventilators | 12 |
| Mild-Moderate | Patients who did not need oxygen administration | 69 |
| COVID-19, coronavirus disease 2019. | | |

### ***Supplementary Table 3:* The sequences of strain used as a reference genome**

| Strain | Complete genome | ORF 1ab polyprotein | Nucleocapsid protein |
| --- | --- | --- | --- |
| COVID-19 Wuhan | NC_045512.2 | YP_009724389.1 | YP_009724397.2 |
| SARS-Coronavirus BJ01 | AY278488 | AAP30028.1 | AAP30037.1 |
| SARS-related Coronavirus | DQ898174 | ABI96956.1 | ABI96968.1 |
| SARS-Coronavirus Tor2 | NC_004718 | NP_828849.7 | YP_009825061.1 |
| MERS-Coronavirus | NC_019843 | YP_009047202.1 | YP_009047211.1 |
| Human β-coronavirus EMC | JX869059 | AFS88944.1 | AFS88943.1 |
| Human β-coronavirus Jordan-N3 | KC776174 | AGH58716.1 | AGH58724.1 |
| Human enteric coronavirus 4408 | FJ415324 | ACJ35483.1 | ACJ35489.1 |
| COVID-19, coronavirus disease 2019; SARS, severe acute respiratory syndrome, MERS, middle-east respiratory syndrome; 3C-like, 3 chymotrypsin-like; ORF, open reading frame. | | | |

### ***Supplementary Figure 1:* The analysis of 3CL^pro^ structure sequence by SV-AUC.**

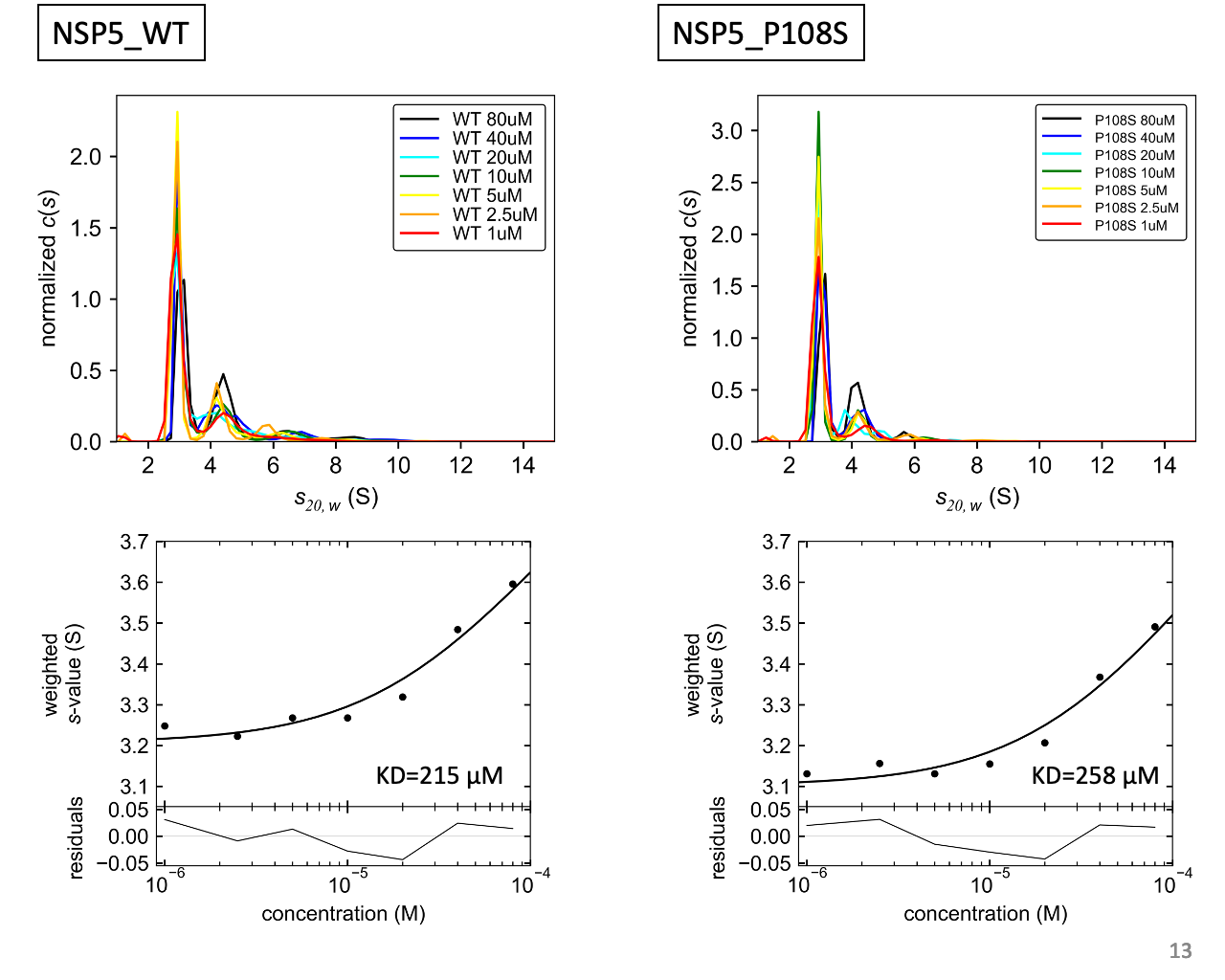

These experiments were performed using the Optima AUC. The collected data were analyzed using continuous c(s) distribution model implemented in program SEDFIT (version 16.2b). The concentration dependence of the weight-average sedimentation coefficient was fitted to the monomer-dimer self-association model implemented in program SEDPHAT (version 15.2b). 3CL^pro^, 3 chymotrypsin-like protease; SV-AUC, sedimentation velocity analytical ultracentrifugation.

### ***Supplementary Figure 2:* The evaluation of chirality between WT and P108S using CD spectroscopy.**

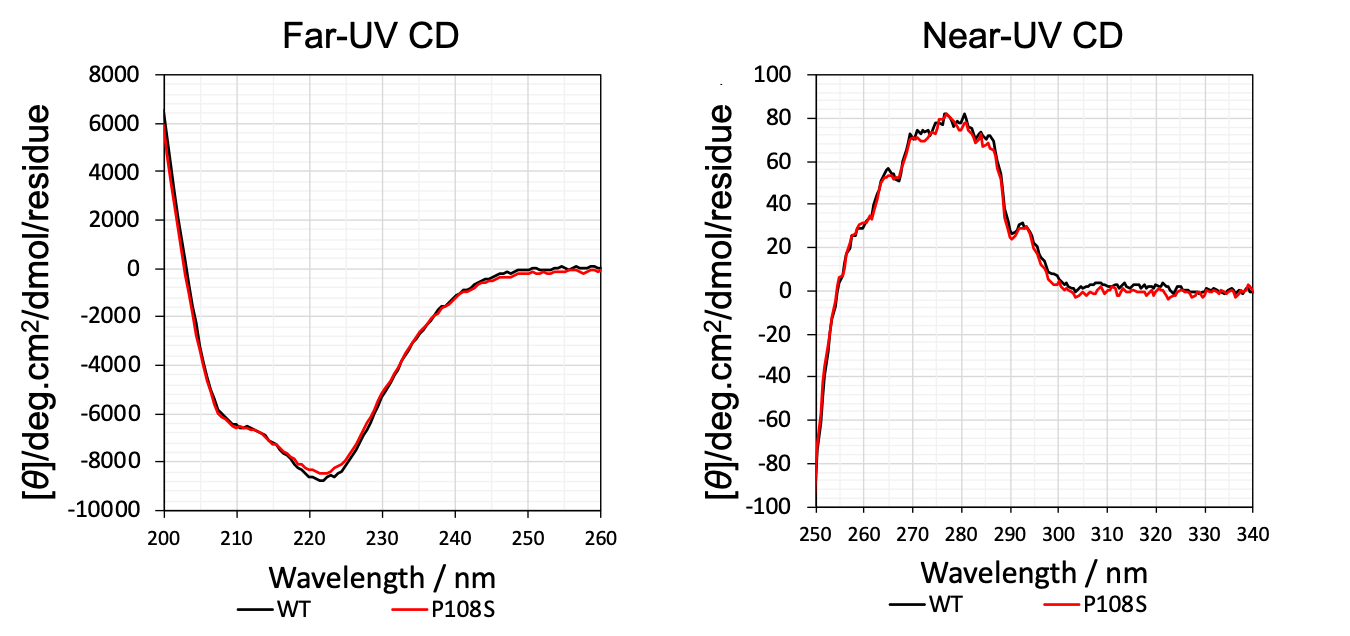

CD spectra were collected in the far-UV (200-260 nm) and the near-UV (250-340 nm) spectral regions. CD, Circular dichroism; UV, ultraviolet; WT, Wuhan-strain type; P108S, Pro108Ser mutant.

### ***Supplementary Figure 3:* The modeled structure and calculated stabilities of Pro151Leu in nucleocapsid protein.**

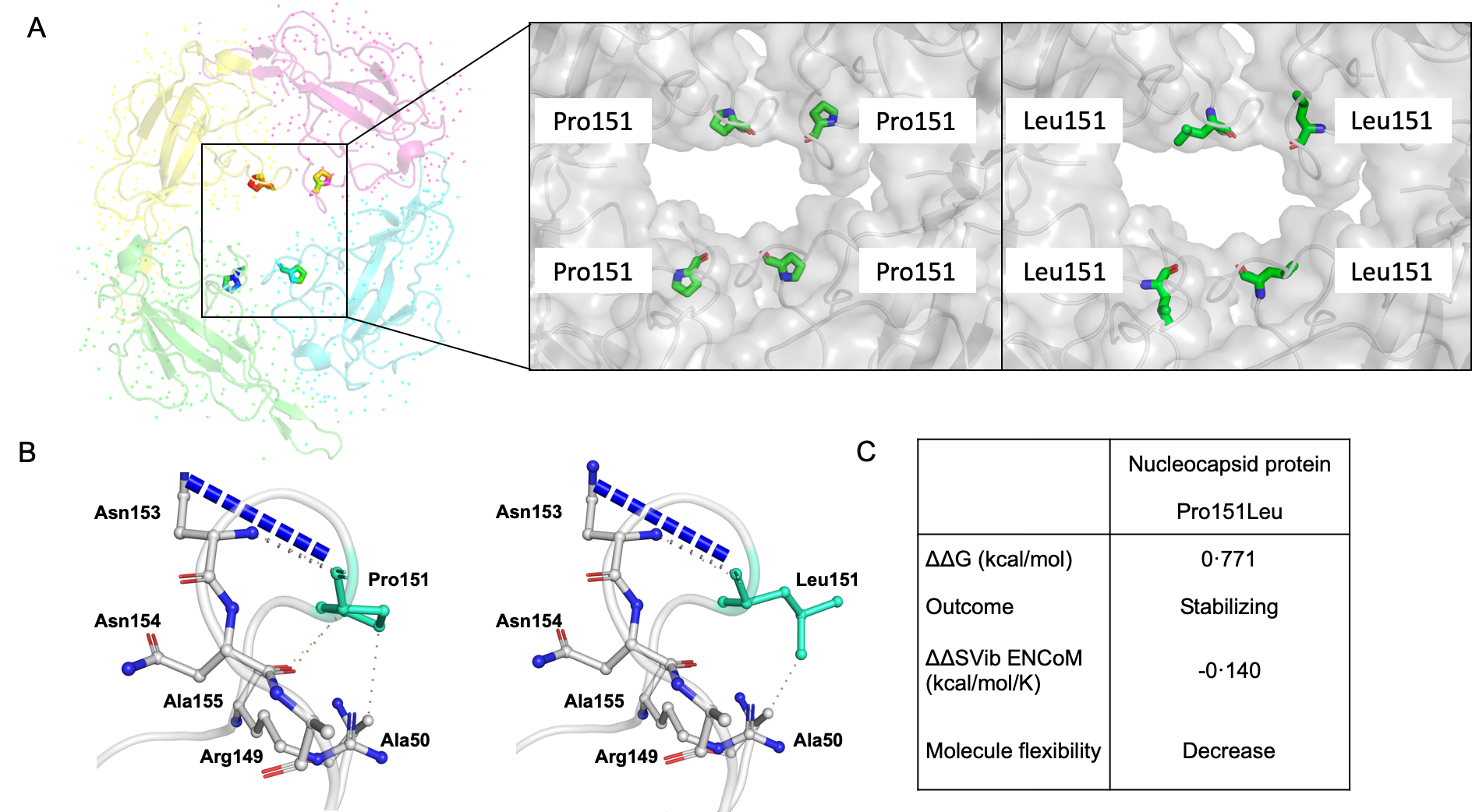

(A) The structure of the tetramer of nucleocapsid protein in SARS-CoV-2 was indicated (PDB ID ‘‘6VYO’’). Pro151Leu mutation is in close proximity to the surface of the tetramer of the critical RNA-binding N-terminal domain. (B) Predicted interatomic interactions and relationship with the surrounding residues between Wuhan-strain type and Pro151Leu mutant in nucleocapsid protein. (C) The impact of Pro151Leu mutation on free energy change in nucleocapsid protein were estimated using DynaMut. SARS-CoV-2, severe acute respiratory syndrome coronavirus 2; PDB, protein data bank; ΔΔ*G*, difference of the free energy change; ΔΔ*S*Vib, difference of the vibrational entropy change between Wuhan-strain type and mutants.

### ***Supplementary Figure 4:* Flowchart of COVID-19 patients.**

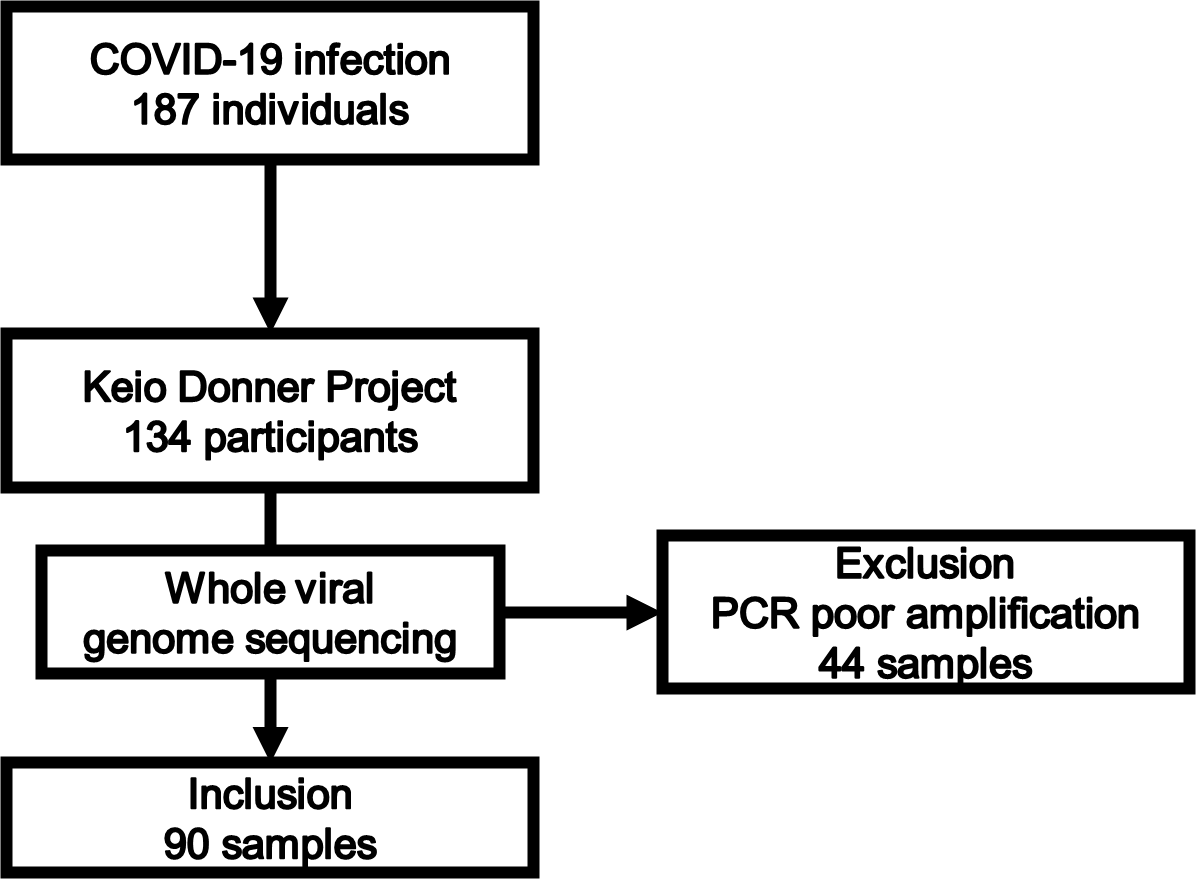

This study finally included 90 out of 187 COVID-19 patients treated during the study period. COVID-19, coronavirus disease 2019; PCR, polymerase chain reaction.
